# Helicobacter pylori fecal antigen positivity in clinically suspected Peptic ulcer disease adults in Dar es Salaam

**DOI:** 10.64898/2026.07.27.26359082

**Authors:** Neema Lweno, Warles Lwabukuna, Alice Gwambegu, Yassin Mgonda

## Abstract

**Background:** *Helicobacter pylori* infection remains a significant public health concern and a leading cause of peptic ulcer disease (PUD). This organism colonizes the gastric mucosa and often remains asymptomatic until complications develop. Despite its global prevalence, the magnitude and association between H. pylori infection and PUD are not well established in many community settings, including Dar es Salaam. This study aimed to determine the fecal prevalence of H. pylori, risk factors, and the correlation between H. pylori and PUD in adults clinically suspected of PUD in Dar es Salaam Communities, 2025.

**Methodology:** A cross-sectional, community-based study was conducted among 390 adults clinically suspected of having PUD in Dar es Salaam over a two-month data-collection period (April-June 2025). Sociodemographic, environmental, behavioral, and clinical data were collected using structured questionnaires, and stool samples were analyzed for *Helicobacter pylori* antigen using standard methods. The primary outcome was Helicobacter pylori infection status, while the secondary outcome was associated risk factors and their correlation with clinical suspicion of PUD. Data were analyzed using SPSS version 25, with categorical variables expressed in frequencies and percentages. Associations were assessed using chi-square tests and multivariable logistic regression, with results reported as adjusted odds ratios and 95% confidence intervals. Spearman’s rank correlation was used to assess the relationship between H.pylori infection and clinical suspicion of PUD. Of the 400 adults enrolled,390(97.5%) provided complete data and were included in the analysis

**Results:** Of 390 participants, 252 (64.6%) were female, and the mean age was 36.9 ± 15.7 years. A total of 161 individuals (41.3%) tested positive for *H. pylori*. Higher Infection rates were observed among adults aged 40–49 years (46.7%), singles (44.6%), those consuming tap water (46.9%), residents of crowded area//s (43.2%), and individuals who consumed alcohol and smoked (50.4% and 44.5%, respectively). Only the source of drinking water showed a significant association with *H. pylori* infection. A positive correlation was observed between *H. pylori* infection and clinically suspected PUD.

**Conclusion:** *H.pylori* infection was common among adults clinically suspected of PUD in the Dar es Salaam communities. The use of tap water for drinking was identified as the most significant risk factor. These findings highlight the need for routine community-based screening and public health interventions to improve water safety and reduce transmission. Future studies should employ longitudinal designs to establish causal relationships between H. pylori infection, behavioral risk factors, and clinical outcomes.

## Introduction

*Helicobacter pylori* is one of the most persistent bacterial infections globally, affecting over 50% of the world’s population and contributing substantially to PUD, chronic gastritis, and gastric cancer.(5). Despite decades of research, the infection continues to exert a considerable burden, especially in low- and middle-income countries where inadequate sanitation, overcrowding, and restricted access to clean water are prevalent. (2),(3). In sub-Saharan Africa, the prevalence of *H. pylori* exceeds 70%, reflecting environmental and behavioral factors that fuel transmission.(6),(7).

In Tanzania, gastrointestinal disorders are among the leading causes of morbidity, with *H. pylori* infection playing a central role.(8). Existing local studies have been hospital-based, relying on patients attending Tanzania care facilities rather than community populations. As a result, there is limited understanding of the true burden, distribution, and determinants of *H. pylori* infection among adults living in the community, particularly those with clinically suspected PUD who may not have access to endoscopic diagnosis.(9). This creates a critical evidence gap that limits effective public health interventions.

Globally, H. pylori infection varies widely from 20–40% in high-income nations to over 70% in developing regions closely linked to sanitation, education, and lifestyle factors.(10),(11),(12). However, few studies have explored community-based fecal antigen testing, a simple, non-invasive diagnostic tool proven sensitive in detecting active infection in low-resource settings such as Uganda and Ethiopia.(13),(14). Unlike serology or invasive biopsy-based tests, stool antigen testing accurately reflects current infection and can guide targeted treatment before complications arise.(13),(15).

Although *H*.*pylori* is implicated in the majority of duodenal and gastric ulcers(1),(16)Not all infected individuals develop PUD, suggesting complex interactions among bacterial virulence, host immunity, and modifiable risk factors such as NSAID use, smoking, and alcohol consumption.(17),(18),(19). Understanding these dynamics within Tanzanian communities is essential for designing targeted interventions.

This study, therefore, aimed to determine the fecal prevalence of *H. pylori*, identify key sociodemographic, behavioral, and environmental risk factors, and assess the correlation between infection and clinical suspicion of PUD among adults in Dar es Salaam. The findings will provide crucial data for designing locally tailored interventions improving diagnostic accuracy, promoting safe water and hygiene practices, and reducing the burden of ulcers and gastric cancer linked to *H. pylori* infection. By shifting focus from hospital settings to the community, this study fills a long-standing evidence gap and strengthens the foundation for preventive gastroenterological health in Tanzania.

The study addressed the gap by generating community-level evidence on H.pylori prevalence, identifying key risk factors, and demonstrating a significant correlation with clinical suspicion of PUD.

### Definition of terms

1. **Clinically suspected peptic ulcer disease** Adults presenting with symptom patterns consistent with peptic ulcer disease based on predefined criteria adapted from the Cornell Medical Index and Dunn’s ulcer pain criteria, including epigastric pain characteristics, meal-related timing, nocturnal pain, symptom relief with food or milk, or a prior clinician-diagnosed ulcer. This definition is symptom-based and does not require endoscopic confirmation.
2. **Helicobacter pylori infection** Presence of *H. pylori* antigen detected in stool samples using a validated fecal (stool) antigen test, indicating active infection.
3. **Fecal (stool) antigen positivity** A positive result on the *H. pylori* stool antigen test cassette, defined by the appearance of both control and test lines according to the manufacturer’s instructions.
4. **Source of drinking water** the primary household water source used for drinking, categorized as tap water, well water, or bottled water, based on participant self-report.
5. Housing conditions were assessed based on the number of individuals sharing sleeping rooms within a household.
  - **Overcrowding** more than three individuals sharing a single sleeping room, consistent with commonly used public health housing standards.
  - **Adequate housing conditions:** Households with three or fewer individuals per sleeping room.
6. **Smoking** Self-reported current use of cigarettes at the time of the study, regardless of frequency or duration.
7. **Alcohol use** Self-reported consumption of alcoholic beverages within the past 12 months, categorized as yes or no.
8. **NSAID use** Self-reported use of non-steroidal anti-inflammatory drugs, categorized as regular, occasional, rare, or never.
9. **Family history of *H. pylori* infection** Self-reported history of a prior medical diagnosis of *H. pylori* infection in a first-degree relative.
10. **Dietary habit:** Predominant pattern of food consumption categorized as balanced diet, fast food–based diet, or poor diet, based on participant responses to structured dietary questions
  - **Balanced diet:** Regular consumption of a mixture of staple carbohydrates, protein sources, fruits, and vegetables.
  - **Fast-food–based diet:** Predominant consumption of commercially prepared foods, including fried foods, processed meals, and street-vended food.
  - **Poor diet:** Irregular meals or diets lacking diversity in major food groups.
11. **Source of daily meals** The primary source from which participants obtained their daily meals, categorized as street-vended food or other sources (home-cooked, hotel, or institutional meals

## Methods

### Study Design

A community-based cross-sectional study was conducted among clinically suspected PUD adults in Dar es Salaam communities.

### Sampling and recruitment techniques

A multistage sampling technique was used to recruit study participants in Dar es Salaam. In the first stage, one district was randomly chosen from the five districts in Dar es Salaam (Kigamboni, Kinondoni, Ilala, Temeke, and Ubungo). In the second stage, a list of administrative divisions within the chosen district was compiled, and one division was selected randomly. In the third stage, two administrative wards were randomly selected from the designated division, resulting in the selection of two streets. In the fourth stage, sub-streets within the selected streets were randomly selected; the 1st, 2^nd,^ and 5th sub-streets were chosen from one street, while the 4th, 5^th,^ and 7th sub-streets were selected from another street. In the fifth stage, households within the selected sub-streets were chosen using systematic random sampling, with every 2nd household on the list included. In each selected household, adults who met the inclusion criteria and provided consent were interviewed, and confidential stool collection was conducted. Those who did not meet the inclusion criteria were excluded, ensuring the required sample size of 374 participants was achieved. The source population comprised adult residents living in selected urban communities within Dar es Salaam, Tanzania. Eligible participants were adults aged 18 years and above with a clinical suspicion of peptic ulcer disease based on Cornell Medical Index criteria, who were able to communicate in Swahili or English and provided informed consent. Participants were enrolled through systematic sampling during household visits.

The recruitment period lasted eight weeks, from April to June 2025. Individuals who had used antibiotics, proton pump inhibitors, or long-term antacids within the preceding four weeks were excluded to preserve the diagnostic accuracy of stool antigen testing.

## Data Collection

### Sociodemographic and Clinical Variables

A structured questionnaire captured sociodemographic data (participants’ age, gender, education level, employment status, and marital status), as well as the Environmental and Living Conditions section (participants’ sources of drinking water, availability of toilet facilities, handwashing practices, and overall living circumstances). The Behavioral Factors included smoking habits, alcohol use, NSAID use, family history of H. pylori infection, which was determined through self-report, based on participants’ recall of a prior medical diagnosis of H. pylori infection in first-degree relatives, dietary habits, and sources of the daily diet. Clinical suspicion of peptic ulcer disease (PUD) was assessed using symptom-based criteria derived from the Cornell Medical Index (CMI) and diagnostic questions based on Dunn’s criteria for ulcer-type pain(20). A total of 11 gastrointestinal symptom questions adapted from the CMI evaluated symptoms such as poor appetite, frequent stomach discomfort, bloating, belching, indigestion, and persistent stomach problems. Responses were recorded as “yes” or “no” and coded as 1 and 0, respectively. In addition, six diagnostic questions were used to characterize ulcer-related pain patterns and medical history. These included whether participants had ever experienced episodes of stomach pain, the timing of pain in relation to meals, whether the pain was relieved by food or milk, whether it awakened them at night, whether a physician had previously diagnosed an ulcer, and whether the diagnosis had been confirmed by esophagogastroduodenoscopy. Clinical suspicion of PUD was determined using three predefined criteria. Criterion 1 identified participants with a classic duodenal ulcer pain pattern, defined as a history of stomach pain occurring two hours or more after eating and relieved by food or milk. Criterion 2 identified participants who reported stomach pain that awakened them at night, a recognized symptom of ulcer disease. Criterion 3 included participants who reported a previous physician diagnosis of ulcer confirmed by esophagogastroduodenoscopy, representing documented disease. Participants meeting any of these criteria were classified as having symptom-defined clinical suspicion of PUD. The full questionnaire and detailed ulcer classification criteria are provided in the Supporting Information (S1 File).

### Laboratory Variables

The H. pylori fecal antigen test was performed using the DPL H. pylori Stool Kit (Nantong EGENS Biotechnology Co., Ltd., Nantong, China; 2024 version). The kit includes all necessary components for collection and testing, including collection tubes, buffer saline, and test cassettes. Participants provided stool samples in clean, labeled disposable containers. A research assistant transferred a portion of each sample into a tube containing Phosphate Buffered Solution (PBS) to homogenize the specimen. Three drops of the homogenized mixture were applied to the antigen detection cassette, and the results were read after 15 minutes. Two lines indicated a positive result, one line a negative result, and invalid tests were repeated according to the manufacturer’s instructions.

### Ethical Approval and consent to participate

Ethical approval was obtained from Kairuki University Institution Research and Ethics Committee (KU/IREC/27.10/553). Permission for data collection was granted by the Dar es Salaam regional administrative secretary (EA.260/307/01B/220). Written informed Consent was obtained from all study participants.

## Data analysis

The data were analyzed using SPSS Version 25 (IBM Corp., Armonk, NY). Chi-square tests assessed associations between variables. Predictors with p ≤0.20 were included in a multivariable logistic regression. Adjusted odds ratios (aOR) with 95% confidence intervals were calculated, with significance set at p<0.05. Model fit was assessed using the Hosmer–Lemeshow goodness-of-fit test, and multicollinearity among independent variables was evaluated using the Variance Inflation Factor (VIF).

## Results

A total of 400 adults were recruited over 2 months (April to June 2025).10 participants had incomplete data (did not produce a stool sample), leaving 390 participants with complete data for analysis.

### The baseline characteristics (social demographics & history) of adults with clinically suspected Peptic ulcer disease in the Dar es Salaam Community

Table 1: Participants had a mean age of 36.9 ± 15.7 years, most were female (64.6%), and aged 18-29 years (43.3%). Nearly half were married (46.4%) and had primary education (47.9%), while most were employed (81.8%). Over half used tap water (57.9%), had poor toilet availability (49.7%), and practiced handwashing (56.4%). Many lived in overcrowded conditions (68.2%). Lifestyle behaviors included smoking (31.8%), alcohol use (72.6%), and regular painkiller consumption (48.2%). A large proportion had a family history of H. pylori (84.6%), consumed fast food (82.8%), and obtained daily meals from street vendors (93.3%).

**Table 1:** The baseline characteristics (social demographics and history) of adults clinically suspected of Peptic ulcer disease in the Dar es Salaam Community. (N=390)

| Variables | Frequency(%) |
| --- | --- |
| **Gender** |  |
| Male | 138(35.4) |
| Female | 252(64.6) |
| **Age** |  |
| 18-29 | 169(43.3) |
| 30-39 | 75 (19.2) |
| 40-49 | 75 (19.2) |
| 50 and > | 71 (18.2) |
| **Education level** |  |
| Not formal | 27( 6.9) |
| Primary | 187(47.9) |
| Secondary | 126(32.3) |
| Higher education | 50(12.8) |
| **Employment status** |  |
| Employed | 319(81.8.) |
| Unemployed | 71(18.2) |
| **Marital Status** |  |
| Single | 92( 23.6) |
| Married | 181(46.4) |
| Divorced | 11( 2.8) |
| Widow/Widower | 32( 8.2) |
| Cohabiting | 74(19.0) |
| **Source of drinking water** |  |
| Tap water | 226(57.9) |
| Well water | 130(33.3) |
| Bottled water | 34(8.7) |
| **Availability of Toilet** |  |
| Excellent | 8(2.1) |
| Good | 175(44.9) |
| Poor | 194(49.7) |
| Very Poor | 13(3.3) |
| **Hand-washing habit** |  |
| Yes | 220(56.4) |
| No | 170(43.6) |
| **Living Environment** |  |
| Adequate | 87(22.3) |
| Overcrowded | 266(68.2) |
| Poor | 37( 9.5) |
| **Cigarette Smoking Status** |  |
| Yes | 124(31.8) |
| No | 266(68.2) |
| **Alcohol Use Status** |  |
| Yes | 283(72.6) |
| No | 107(27.4) |
| **NSAIDs use** |  |
| Regular | 188(48.2) |
| Occasional | 146(37.4) |
| Rarely | 53(13.6) |
| Never | 3(0.8) |
| **Family h/o of *H. pylori*** |  |
| Yes | 330(84.6) |
| No | 60(15.4) |
| **Dietary habit** |  |
| Poor | 28(7.2) |
| Balanced Diet | 39(10.0) |
| Fast Food | 323(82.8) |
| **Source of the daily diet** |  |
| Street vendors | 364(93.3) |
| Others(home cooked,hotel,institution) | 26( 6.7) |

### The Prevalence of *H. pylori* Infection in Clinically Suspected Peptic Ulcer Disease Adults in Dar es Salaam Community

H. pylori prevalence was 41.3% (161/390 participants were positive)

### The distribution of social demographic characteristics (Gender, Age, Education level, Employment status, and marital status) of H. pylori infection in adults clinically suspected of Peptic ulcer disease in the Dar es Salaam Community

Table 2 shows that infection was more common in males (p < 0.05). Higher prevalence also appeared among participants aged 40-49 years (46.7%), the employed (42.6%), those with secondary education (46.0%), and singles (44.6%), though these were not statistically significant.

**Table 2:**
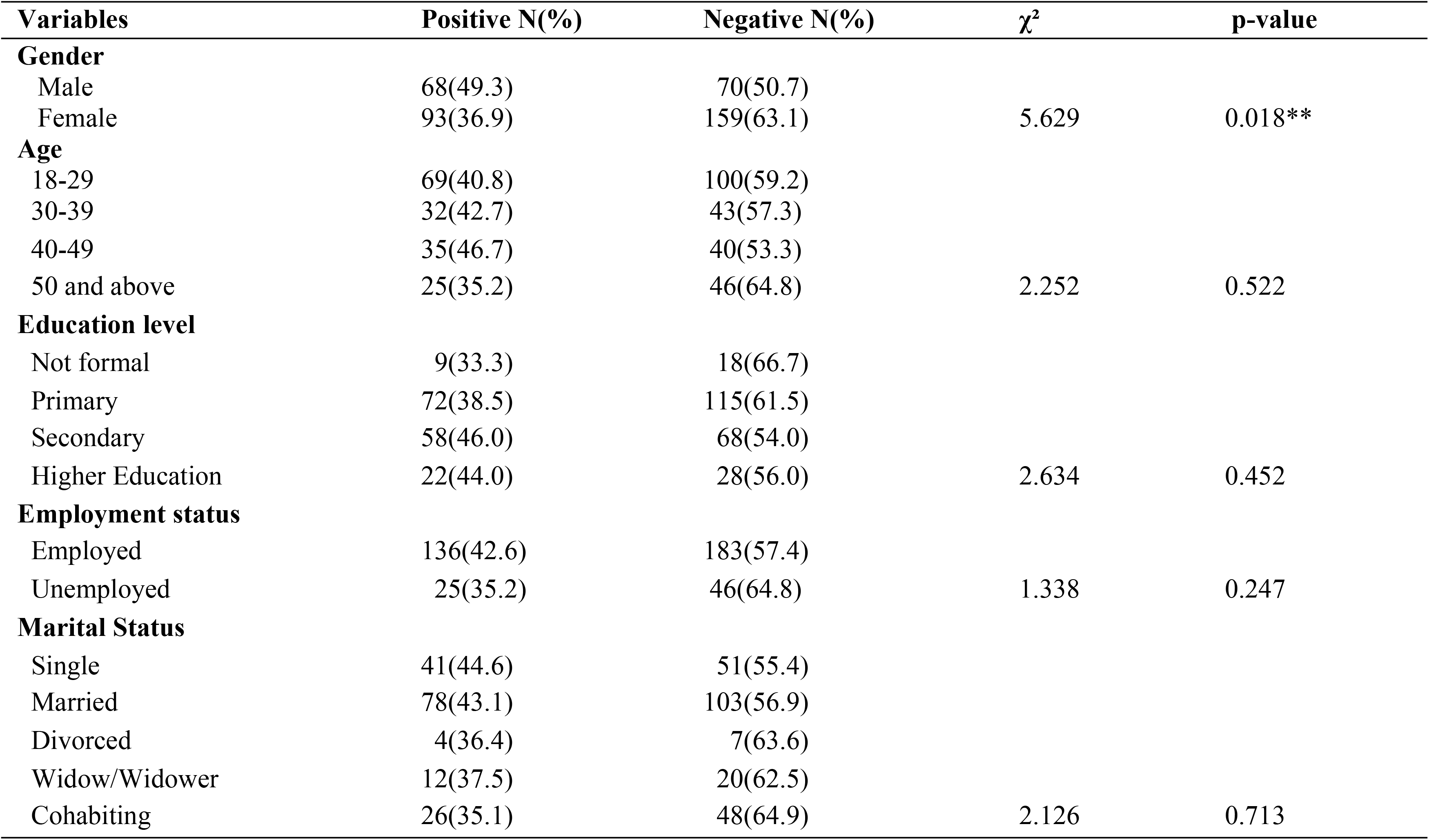
Socio-demographic risk factors of H. pylori infection in adults clinically suspected of Peptic ulcer disease, Dar es Salaam Community. (N=390)

| Variables | Positive N(%) | Negative N(%) | $\chi^2$ | p-value |
| --- | --- | --- | --- | --- |
| **Gender** |  |  |  |  |
| Male | 68(49.3) | 70(50.7) | 5.629 | 0.018** |
| Female | 93(36.9) | 159(63.1) |  |  |
| **Age** |  |  |  |  |
| 18-29 | 69(40.8) | 100(59.2) | 2.252 | 0.522 |
| 30-39 | 32(42.7) | 43(57.3) |  |  |
| 40-49 | 35(46.7) | 40(53.3) |  |  |
| 50 and above | 25(35.2) | 46(64.8) |  |  |
| **Education level** |  |  |  |  |
| Not formal | 9(33.3) | 18(66.7) | 2.634 | 0.452 |
| Primary | 72(38.5) | 115(61.5) |  |  |
| Secondary | 58(46.0) | 68(54.0) |  |  |
| Higher Education | 22(44.0) | 28(56.0) |  |  |
| **Employment status** |  |  |  |  |
| Employed | 136(42.6) | 183(57.4) | 1.338 | 0.247 |
| Unemployed | 25(35.2) | 46(64.8) |  |  |
| **Marital Status** |  |  |  |  |
| Single | 41(44.6) | 51(55.4) | 2.126 | 0.713 |
| Married | 78(43.1) | 103(56.9) |  |  |
| Divorced | 4(36.4) | 7(63.6) |  |  |
| Widow/Widower | 12(37.5) | 20(62.5) |  |  |
| Cohabiting | 26(35.1) | 48(64.9) |  |  |

### The Distribution of Environmental Risk Factors(Source of water, Availability of Toilet, Hand washing habit, and living environment) of H. pylori infection in clinically suspected Peptic ulcer disease adults in Dar es Salaam Community

Environmental risk factors (Table 3) showed significantly higher infection among tap water users (46.9%), smokers (50.4%), and alcohol users (44.5%) (all p < 0.05). Higher, though non-significant, rates occurred among participants in poor living conditions (43.2%), those practicing handwashing (42.9%), individuals with a family history of H. pylori (43.0%), fast-food consumers (43.3%), rare NSAID users (50.9%), and those dependent on street vendors for meals (41.8%). The lowest rate (12.5%) was among those with excellent toilet facilities.

**Table 3:** Environmental Risk Factors of H. pylori infection in clinically suspected Peptic ulcer disease adults in Dar es Salaam Community. (N=390)

| Variables | Positive N(%) | Negative N(%) | $\chi^2$ | p-value |
| --- | --- | --- | --- | --- |
| **Source of water** |  |  |  |  |
| Tap water | 106(46.9) | 120(53.1) | 7.486 | 0.024** |
| Well water | 42(32.3) | 88(67.7) |  |  |
| Bottled water | 13(38.2) | 21(61.8) |  |  |
| **Availability of Toilet** |  |  |  |  |
| Excellent | 1(12.5) | 7(87.5) | 4.364 | 0.225 |
| Good | 77(44.0) | 98(56.0) |  |  |
| Poor | 79(40.7) | 115(59.3) |  |  |
| Very Poor | 4(30.8) | 9(69.2) |  |  |
| **Hand-washing habit** |  |  |  |  |
| Yes | 94(42.9) | 125(57.1) | 0.554 | 0.457 |
| No | 67(39.2) | 104(60.8) |  |  |
| **Living environment** |  |  |  |  |
| Adequate | 33(37.9) | 54(62.1) | 0.536 | 0.765 |
| Overcrowded | 112(42.1) | 154(57.9) |  |  |
| Poor | 16(43.2) | 21(56.8) |  |  |

### The Distribution of Behavioral Risk Factors(Cigarette smoking, Alcohol use, NSAIDs use, Family history of *H. pylori*, Dietary habit, and Source of daily diet) of H. pylori Infection in Clinically Suspected PUD Adults in Dar es Salaam Community

Behavioral risk factors (Table 4) showed significantly higher H. pylori infection rates among smokers (50.4%) and alcohol users (44.5%) (both p < 0.05). Higher, though non-significant, infection rates were observed among participants with a family history of H. pylori (43.0%), fast-food consumers (43.3%), participants who rarely used NSAIDs (50.9%), and those who relied on street-vended meals (41.8%). The lowest infection rate was seen among participants consuming a balanced diet (30.8%).

**Table 4:**
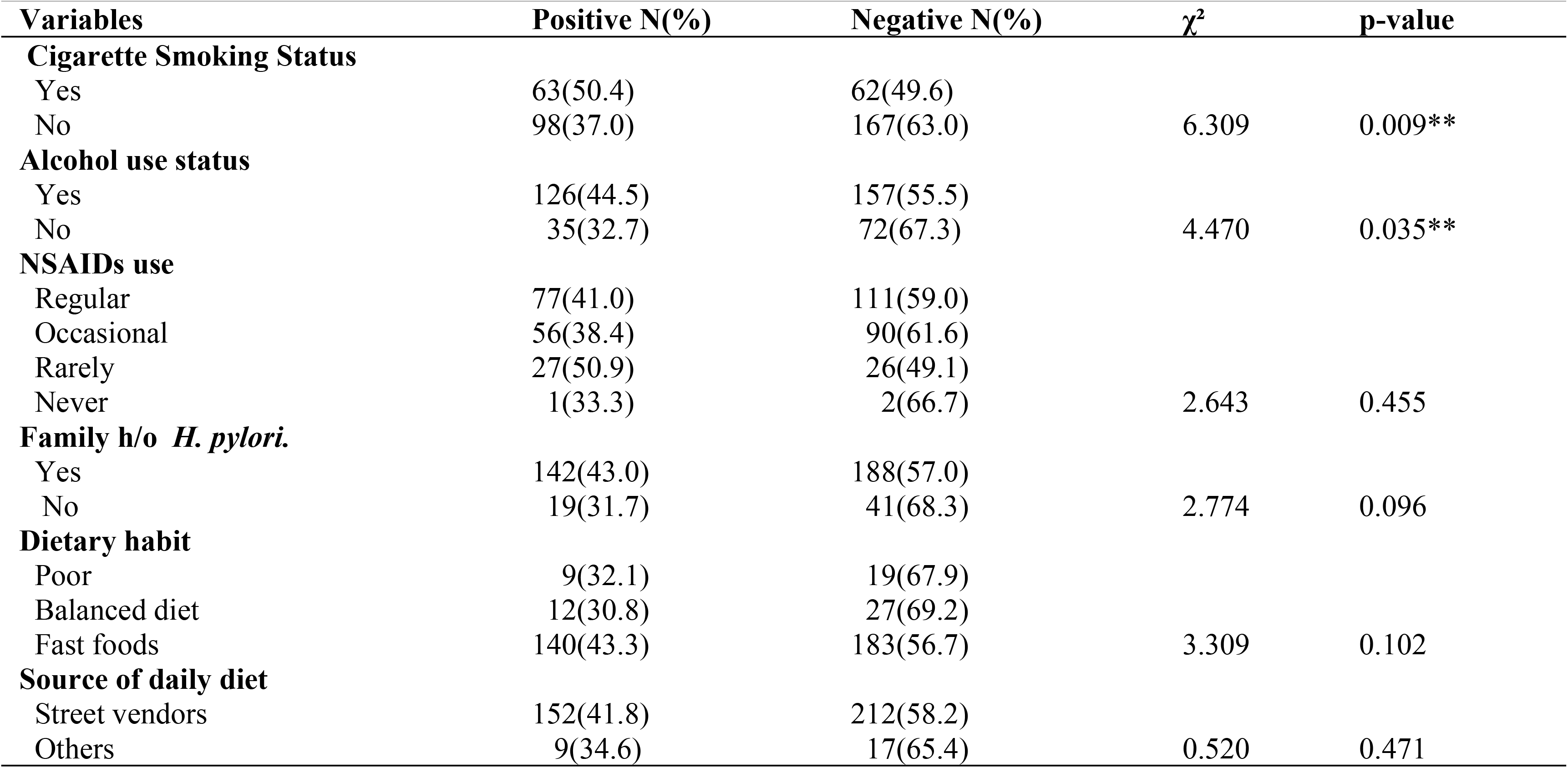
Behavioral Risk Factors of H. pylori infection in clinically suspected Peptic ulcer disease adults in Dar es Salaam Community. (N=390)

| Variables | Positive N(%) | Negative N(%) | $\chi^2$ | p-value |
| --- | --- | --- | --- | --- |
| **Cigarette Smoking Status** |  |  |  |  |
| Yes | 63(50.4) | 62(49.6) | 6.309 | 0.009** |
| No | 98(37.0) | 167(63.0) |  |  |
| **Alcohol use status** |  |  |  |  |
| Yes | 126(44.5) | 157(55.5) | 4.470 | 0.035** |
| No | 35(32.7) | 72(67.3) |  |  |
| **NSAIDs use** |  |  |  |  |
| Regular | 77(41.0) | 111(59.0) | 2.643 | 0.455 |
| Occasional | 56(38.4) | 90(61.6) |  |  |
| Rarely | 27(50.9) | 26(49.1) |  |  |
| Never | 1(33.3) | 2(66.7) |  |  |
| **Family h/o *H. pylori*.** |  |  |  |  |
| Yes | 142(43.0) | 188(57.0) | 2.774 | 0.096 |
| No | 19(31.7) | 41(68.3) |  |  |
| **Dietary habit** |  |  |  |  |
| Poor | 9(32.1) | 19(67.9) | 3.309 | 0.102 |
| Balanced diet | 12(30.8) | 27(69.2) |  |  |
| Fast foods | 140(43.3) | 183(56.7) |  |  |
| **Source of daily diet** |  |  |  |  |
| Street vendors | 152(41.8) | 212(58.2) | 0.520 | 0.471 |
| Others | 9(34.6) | 17(65.4) |  |  |

### Factors associated with Helicobacter Pylori infection in clinically suspected PUD adults, in the Dar es Salaam Community

Multivariable analysis(Table 5) identified drinking water source as the only independent predictor of infection (p < 0.05), with well-water users being significantly less likely to be infected.

**Table 5:** The multivariate logistic regression of risk factors of *H*.*pylori* infection in clinically suspected Peptic ulcer disease adults in Dar es Salaam Community. (N=390)

| Variables | UOR for 95% C.I (lower-upper) | P-value | AOR FOR 95% C.I (lower-Upper) | P-value |
| --- | --- | --- | --- | --- |
| **Gender** |  |  |  |  |
| Male(ref) |  |  |  |  |
| Female | 1.661(1.091-2.529) | 0.018 | 1.182(0.549-2.544) | 0.670 |
| **Source of water** |  |  |  |  |
| Tap water(ref) |  |  |  |  |
| Well water | 0.566(0.364-0.879) | 0.011 | 0.574(0.362-0.911) | 0.018** |
| Bottled water | 0.870(0.420-1.793) | 0.706 | 0.780(0.363-1.672) | 0.522 |
| **Cigarette Smoking Status** |  |  |  |  |
| Yes | 1.770(1.150-2.725) | 0.009 | 1.279(0.563-2.904) | 0.556 |
| No(ref) |  |  |  |  |
| **Alcohol use status** |  |  |  |  |
| Yes | 1.651(1.035-2.634) | 0.035 | 1.318(0.778-2.232) | 0.304 |
| No(ref) |  |  |  |  |
| **Dietary Habits** |  |  |  |  |
| Poor(ref) |  |  |  |  |
| Balanced diet | 0.603(0.296-1.228) | 0.163 | 0.944(0.323-2.759) | 0.916 |
| Fast foods | 1.676(0.956-2.937) | 0.071 | 1.376(0.590-3.210) | 0.460 |
| **Family h/o *H. pylori*** |  |  |  |  |
| Yes(ref) |  |  |  |  |
| No | 1.630(0.907-2.928) | 0.096 | 1.417(0.767-2.619) | 0.266 |

### The Correlation between *H. Pylori* infection and clinical suspicion of PUD among adults in the Dar es Salaam Community

Infection was most frequent among participants meeting criterion 1 (42.2%), followed by criterion 2 (38.1%) and criterion 3 (14.3%). Spearman correlation analysis (Table 6) demonstrated a strong positive association across all criteria (ρ = 0.634, 0.933, and 0.884, respectively; p < 0.05 for all). These results reflect an association between H. pylori infection and symptom-defined clinical suspicion, but do not indicate confirmed PUD diagnoses, as the criteria are based on symptoms that may overlap with other gastrointestinal conditions

**Table 6:** The Correlation between H. Pylori infection and clinical suspicion of Peptic ulcer disease(Criteria 1-3) among adults in the Dar es Salaam Community. N= 390.

| Criteria | Positive, N(%) | Negative,N(%) | Spearman's rank ($\rho$) | P-value |
| --- | --- | --- | --- | --- |
| Criterion 1 | 144(42.2) | 197(57.8) | 0.634 | 0.001*** |
| Criterion 2 | 16(38.1) | 26(61.9) | 0.933 | 0.000*** |
| Criterion 3 | 1(14.3) | 6(85.7) | 0.884 | 0.000*** |

## Discussion

This study found a 41.3% prevalence of H.pylori infection among adults clinically suspected of having peptic ulcer disease in Dar es Salaam communities. The source of drinking water was the only independent predictor of infection, with tap water users at higher risk. Smoking and alcohol use were associated with infection in univariate analyses but were not significant in multivariable regression models. Family history of H.pylori and consumption of fast food showed higher, yet non-significant, prevalence. A strong correlation was observed between H.pylori infection and clinical suspicion of PUD, supporting the reliability of symptom-based screening in resource-limited settings.

The prevalence observed aligns with studies from Northern Tanzania(9), suggesting that *H. pylori* infection remains common in urban and semi-urban populations. The association with drinking water reflects the fecal-oral transmission route, particularly in areas with limited sanitation and water treatment. These findings are consistent with reports from other East African countries, reinforcing the importance of environmental factors in *H. pylori* epidemiology.(21). The lack of independent associations with smoking, alcohol, or dietary factors contrasts with some regional meta-analyses.(21), possibly due to differences in behavioral definitions, population characteristics, or exposure levels.

Our findings also highlight the role of household and community-level factors. Although family history and fast-food consumption were not significant predictors, shared living spaces, limited hygiene, and poor food handling may contribute to clustering of infections. Reliance on street-vended food is common in urban Dar es Salaam due to affordability, work schedules, and limited access to home-prepared meals, particularly among employed adults. This context helps interpret the high prevalence observed. Conversely, consumption of balanced diets may support gastric health and reduce infection risk, consistent with previous studies showing protective effects of nutrient-rich foods.(14).

A strong association was observed between *H. pylori* infection and clinical suspicion of PUD, consistent with findings from Nigeria. In resource-limited settings, restricted access to diagnostic tools may allow infections to progress to symptomatic disease, which likely explains the high concordance between infection status and PUD-related symptoms (20).

The study’s strengths include the use of a validated symptom assessment tool, a reliable noninvasive diagnostic method, and multivariable analysis to identify independent risk factors. Limitations include reliance on self-reported behaviors and symptoms, which may introduce recall or reporting bias, and the cross-sectional design, which precludes causal inference. Additionally, the higher proportion of females in our sample than males reflects greater healthcare-seeking behavior among women and higher availability of women during daytime community-based recruitment, rather than the underlying population gender distribution, which should be considered when interpreting the findings.

These results are generalizable to urban Tanzanian adults and similar resource-limited settings where water and sanitation challenges persist. Future research should evaluate interventions to improve water quality, community-level screening programs, and longitudinal studies to establish causal relationships between H. pylori infection, behavioral risk factors, and clinical outcomes.

In conclusion, *Helicobacter pylori* infection was common among adults with symptom-defined clinical suspicion of peptic ulcer disease in Dar es Salaam. Drinking water source was the only independent predictor of infection, highlighting the potential role of water safety in reducing transmission. In resource-limited settings where access to endoscopy and advanced diagnostic tools is limited, symptom-based clinical assessment combined with non-invasive *H. pylori* stool antigen testing may provide a feasible approach for identifying individuals at risk of peptic ulcer disease. These findings suggest that validated symptom assessment tools could support targeted testing strategies in community settings. However, further studies are needed to validate this approach and to determine whether it can improve patient outcomes while minimizing unnecessary antibiotic use.

**Figure 1:**
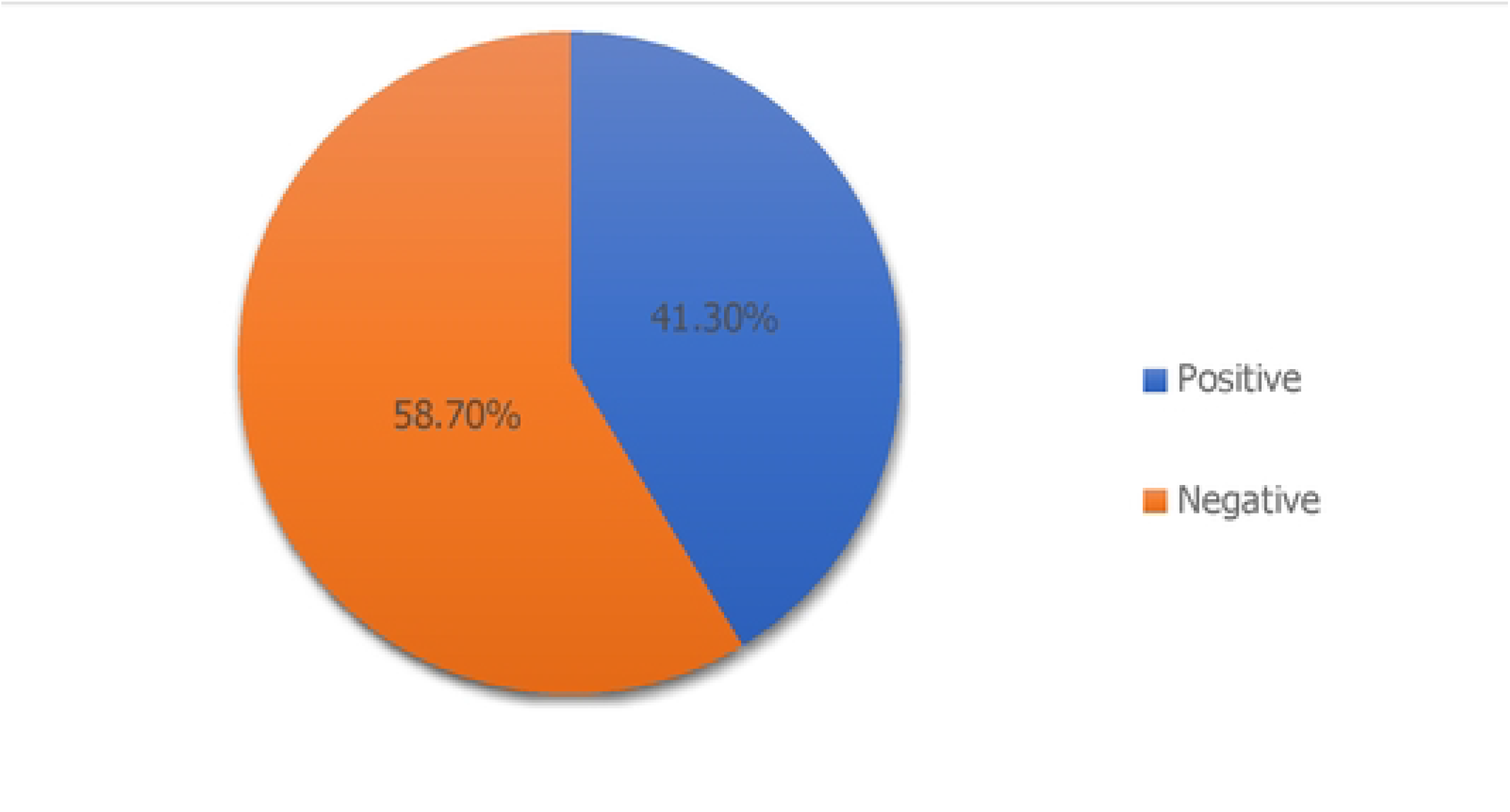
The Prevalenceof H. pylori infection among clinically suspected Peptic ulcer disease adults in the Dar es Salaan1 Comn1unity. (N=390).

## Data Availability

All relevant data underlying the findings of this study are provided within the manuscript, Supporting Information files, and the accompanying de-identified minimal dataset submitted with this manuscript.

## Declarations

### Consent for publications

Not applicable.

### Availability of data and materials

The de-identified minimal dataset underlying the findings of this study is provided as Supporting Information (S1 Dataset). All other relevant data are within the manuscript and its Supporting Information files.

### Authors’ contributions

All authors contributed equally to conception and research proposal development. NL contributed to data collection, entry, and analysis. WL, AG, and YM were instrumental in statistical analysis. All authors read and gave final approval for the manuscript’s submission and publication.

## Acknowledgements

Appreciation is extended to Kairuki University, research assistants, and study participants.

## Supporting information

### S1 File: Clinical questionnaire used to assess suspicion of peptic ulcer disease

The questionnaire, adapted from the Cornell Medical Index and Dunn’s ulcer pain criteria used to assess symptoms suggestive of peptic ulcer disease(20).

Have you experienced any of the following symptoms? *(Check all that apply*.)

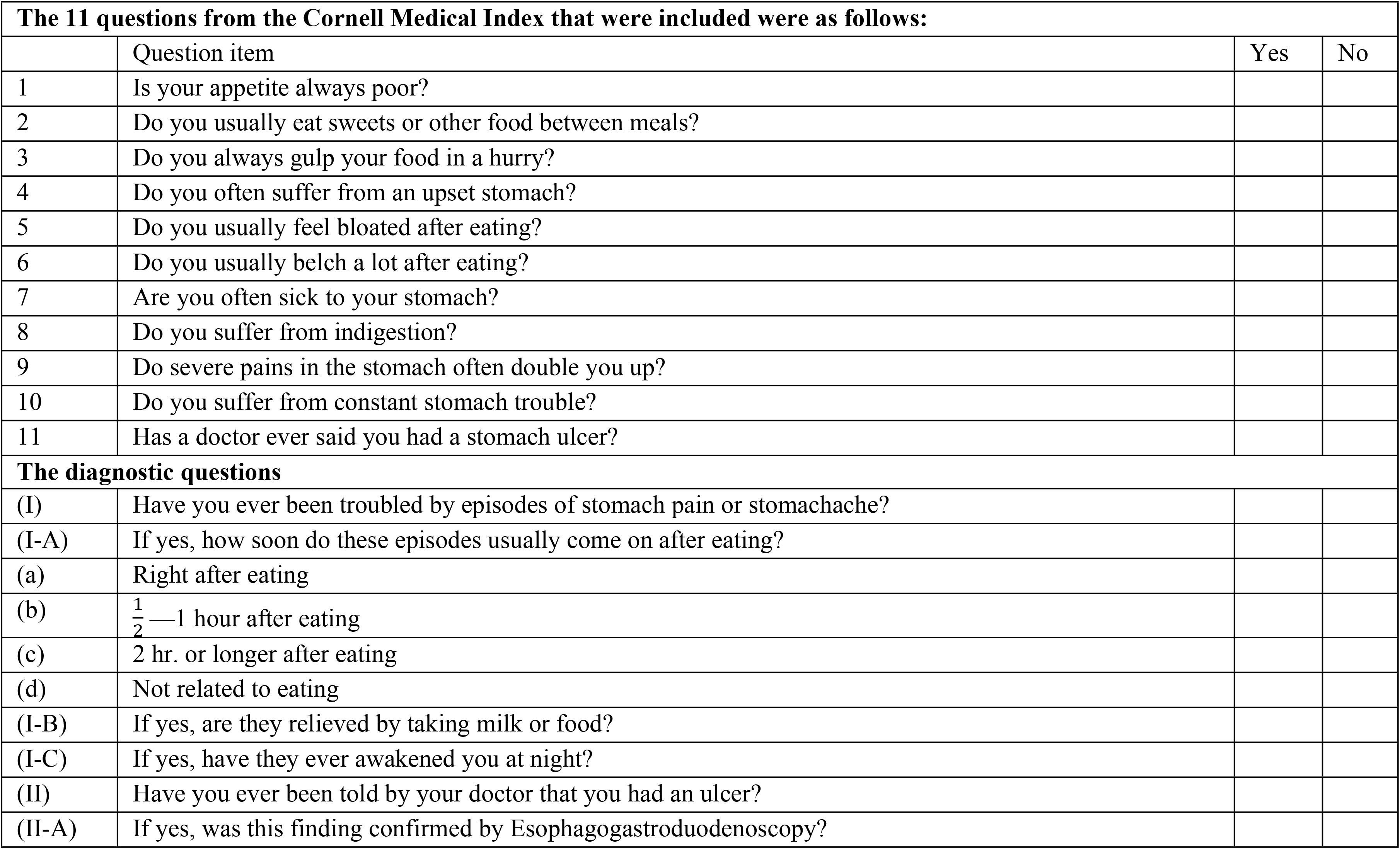

## Ulcer Classification

- **Criteria 1: Positive answer to:**
  ∘ Question I: Have you ever been troubled by stomach pain or aches?
  ∘ And Question I-A-c: How soon do these episodes usually come on after eating? (2 hours or longer)
  ∘ And Question I-B: Are they relieved by taking milk or food?
- **Criteria 2: Positive answer to:**
  ∘ Question I: Have you ever been troubled by stomach pain or aches?
  ∘ And Question I-C: Have they ever awakened you at night?
- **Criteria 3: Positive answer to:**
  ∘ Question II: Have you ever been told by your doctor that you had an ulcer?
  ∘ And Question II-A: Was this finding confirmed by Esophagogastroduodenoscopy?

